# Predicting inpatient flow at a major hospital using interpretable analytics

**DOI:** 10.1101/2020.05.12.20098848

**Authors:** Dimitris Bertsimas, Jean Pauphilet, Jennifer Stevens, Manu Tandon

## Abstract

**Problem definition:** Turn raw data from Electronic Health Records into accurate predictions on patient flows and inform daily decision-making at a major hospital.

**Practical Relevance:** In a hospital environment under increasing financial and operational stress, forecasts on patient demand patterns could help match capacity and demand and improve hospital operations.

**Methodology:** We use data from 63, 432 admissions at a large academic hospital (50.0% female, median age 64 years old, median length-of-stay 3.12 days). We construct an expertise-driven patient representation on top of their EHR data and apply a broad class of machine learning methods to predict several aspects of patient flows.

**Results:** With a unique patient representation, we estimate short-term discharges, identify long-stay patients, predict discharge destination and anticipate flows in and out of intensive care units with accuracy in the 80%+ range. More importantly, we implement this machine learning pipeline into the EHR system of the hospital and construct prediction-informed dashboards to support daily bed placement decisions.

**Managerial Implications:** Our study demonstrates that interpretable machine learning techniques combined with EHR data can be used to provide visibility on patient flows. Our approach provides an alternative to deep learning techniques which is equally accurate, interpretable, frugal in data and computational power, and production-ready.

## 1 Introduction

In a hospital environment under increasing financial and operational stress, improvement in care delivery requires “the utilization of advanced data analytics to […] forecast patient demand patterns, and match capacity and demand” (Rutherford et al. 2017). In this regard, combining patient-level information from Electronic Health Records (EHRs) with sophisticated predictive analytics can provide welcome visibility on patient flows and inform hospital operations. Such is the contribution of the present paper: We build an end-to-end machine learning pipeline and convert raw clinical data from EHRs into a unique patient representation. Using this rich set of covariates, we predict operationally relevant outcomes using interpretable machine learning techniques, thus providing insights and explanations alongside highly accurate predictions. With practical impact in mind, we fully integrate our models into the EHR system of a large academic hospital and design user-friendly dashboards to support daily decision-making.

### 1.1 Related work

#### Patient representation

Despite the richness and increasing availability of data in health-care, predictive models are not widely deployed in practice, due to the need to create custom dataset with specific variables for each predictive task. To address this issue, Nguyen et al. (2016), Miotto et al. (2016), Rajkomar et al. (2018) proposed automatized patient representation strategies which analyze EHRs and construct relevant features in an unsupervised way using autoencoder neural networks. Since these approaches do not require an expert to manually define features, they are allegedly more scalable. Surprisingly, however, and to the best of our knowledge, none of these approaches has been integrated within an EHR system of a real-world hospital despite their excellent predictive power on retrospective studies, including the most recent one (Rajkomar et al. 2018). In our opinion, they undermined three major implementation bottlenecks. First of all, the black-box nature of deep learning models impedes adoption from doctors and caregivers which are not engaged in the modeling process. Automation alone does not guarantee scalable implementation. In our experience, involving stakeholders at each point in the process is instrumental in building trust between clinical and analytical teams and deploying the predictive models in production. Secondly, deep learning approaches are extremely expensive in terms of data, human and computing resources, and environmental costs (Strubell et al. 2019). Finally, convolutional and recurrent neural network are excellent at handling unstructured data such as medical notes. However, in practice, notes are rarely available in real-time and raise data privacy issues, especially if third-party computational resources are needed. Consequently, we believe they are better suited for retrospective clinical studies than production-ready real-time analytics.

#### Patient flow prediction

In this work, we focus our attention on inpatients, namely patients who are admitted at the hospital and occupy a bed in an inpatient unit. For this population, patient flows can be divided into two categories: flows out of the hospital, i.e., discharges, and flows between units of the hospital.

At a hospital level, a collection of work (Kim et al. 2014, Zhu et al. 2015, Luo et al. 2017, McCoy et al. 2018) applied time-series methods to predict daily discharge volume. At a patient level, predicting discharge is associated with predicting length of stay. Being a surrogate for negative clinical outcomes as well as operational performance, length of stay has received a vivid interest in the academic literature, often in combination with hospital mortality (see Awad et al. 2017, for a comprehensive rewiew). From a clinical perspective, prolonged length of stay is associated with negative outcomes for the patient, such as increased infection risks. From an operational perspective, patient discharges drive bed availability, which is one of the most critical hospital resources. Thus, accurate length of stay prediction of inpatients could improve care delivery at a patient level, by highlighting current discharge barriers or identifying complex cases, and at a healthcare facility level, through improved resource management and planning. Discharge destination, i.e., where the patient will be discharged to, is another important component of the discharge process. Indeed, further case management resources should be allocated to patients requesting a bed in extended care facilities, while high-mortality-risk patients should be detected early on by the clinicians. We compiled a list of comparable work from the literature in Table 1.

**Table 1:** Condensed literature review. We selected papers which (a) studied similar outcomes, (b) put emphasis on prediction rather than causation, (c) were published recently.

| Reference | Scope | Predicted outcome | Methodology |
| --- | --- | --- | --- |
| Tabak et al. (2014) | All inpatients | Mortality | Linear regression |
| Van Walraven and Forster (2017) | All inpatients | Discharge volume | Survival analysis |
| McCoy et al. (2018) | Hospital level | Discharge volume | Time series |
| Rajkomar et al. (2018) | All inpatients | Mortality, overall LOS > 7 days | Deep learning |
| Safavi et al. (2019) | Surgical inpatients | Remaining LOS < 1 day | Deep learning |

Regarding patient flows between units, the most critical ones are flows to and out of intensive care units (ICUs), for ICUs are expensive and limited resources needed by the most severe patients. Patients who cannot be admitted to an ICU due to congestion have to be admitted to a general care bed, leading to increased length of stay and readmission risk (Kim et al. 2016). Congested ICUs also increase waiting time and congestion in the rest of the hospital such as in the emergency department (Mathews et al. 2018) or regular inpatient wards (Long and Mathews 2018). Troy and Rosenberg (2009), Angelo et al. (2017) incorporates estimates of overall demand for ICU into a simulation framework to inform strategic capacity sizing decisions. In empirical studies, it is not unusual to divide patients depending on whether they need an ICU or not. However, in practice, a patient’s need varies along her stay. In this work, we investigate the question of estimating the probability for each patient to need an ICU bed in the next 24 hours. To the best of our knowledge, this specific question has not been studied in the literature yet.

### 1.2 Contributions and structure

Our contributions can be summarized as follows:

- We propose a simple expertise-driven patient representation framework to capture the state of each inpatient as she stays in the hospital, competitive with the deep learning approaches recently proposed in the literature (Nguyen et al. 2016, Miotto et al. 2016, Rajkomar et al. 2018). Compared to previous work, we use a hospital-centric rather than patient-centric time scale and only leverage features which are reliably available after admission, on a daily basis. Consequently, we successfully integrate our patient representation into a real-world EHR system and now process the data of a 600-bed hospital daily.
- From this unique set of features, we apply a broad collection of machine learning techniques to address four length of stay-related tasks: identify same-day and next-day discharges and predict more-than-7 and more-than-14-day stays. We then investigate the question of predicting discharge destination among home, home with services, extended care facility and death. We also predict the probability for a given patient to need an intensive care bed in the next 24 hours. For all tasks, we match or surpass state-of-the-art methods, even without using raw medical notes. Ensemble methods are the most accurate, but linear models and decision trees provide very good predictive power, together with actionable insights to practitioners thanks to their interpretability.
- While the machine learning methods we apply are not novel, we would like to emphasize the fact that their practical implementation in a real-world context still raises nontrivial questions, to which we provide a less theoretical but more pragmatic answer. For instance, we extensively discuss how to split the data into training and validation sets (Section 2.5) or how to convert individual risk scores (between 0 and 1) into binary predictions and hospital-level estimates (Appendix D).
- Our work illustrates that emphasis on modeling and interpretability does not hinder predictive accuracy nor scalability. On the contrary, the variety of predictive tasks we cover, together with high level of accuracy, demonstrates that an expertise-driven patient representation framework can be equally powerful and versatile as neural network approaches. In addition, it leads to more interpretable features, achieves higher engagement from the clinicians and care providers, and requires less data and computational resources. As a result, we were able to conduct the project from initial data exploration to production-level deployment in less than twelve months.
- Finally, we empirically evaluate the operational impact of the deployment of our tool on hospital operations. In particular, we investigate the impact on patient admissions from the Emergency Department. Among others, a difference-in-differences analysis reveals a 4% reduction (*p*-value < 0.01) in off-service placements thanks to our machine learning-informed dashboard.

The rest of the paper is organized as follows: In Section 2, we present the data and outcomes of interest, describe the patient representation we designed with physicians and outline our data analysis methodology. In Section 3, we report and compare the predictive power of five machine learning techniques on retrospective data. More importantly, we implement our modeling framework and machine learning algorithms into the EHR system of a large academic medical center. We discuss implementation issues, operational benefits, and empirical performance in production in Section 4.

## 2 Data description and methodology

In this section, we describe the data, our patient representation and the machine learning methodology we applied for our retrospective study.

### 2.1 Study population

We gather data from the EHRs of inpatients admitted at BIDMC between January 2017 and August 2018. We excluded patients admitted into psychiatry, obstetrics and newborns, as well as observation patients who did not stay overnight. The final cohort consists of 63, 432 unique admissions (41, 726 unique patients), whose demographics and relevant variables are summarized in Table 2. The dataset contains patient demographics, provider orders, ICD10 diagnosis codes from previous and current admissions, medications, blood laboratory values, vital signs and key scores (e.g., pain scale, mobility score). Institutional review board at BIDMC approved the study with waiver of informed consent.

**Table 2:** Summary statistics for the study population in the training and validation (Jan 2017-Apr 2018) and test (May 2018 - Aug 2018) data set.

| | Training and validation<br>($n = 50,467$) | Test<br>($n = 12,965$) |
| --- | --- | --- |
| *Demographics* |  |  |
| Age, median (IQR), years | 64 (23) | 65 (24) |
| Female sex, no. (%) | 25,196 (49.9%) | 6,537 (50.4%) |
| Homeless, no. (%) | 54 (0.1%) | 7 (0.1%) |
| Not english speaking, no. (%) | 5,066 (10.0%) | 1,340 (10.3%) |
| Body mass index, median (IQR) | 27.40 (8.52) | 27.40 (8.52) |
| Pre-existing comorbidities, median (IQR) | 0 (5) | 0 (6) |
| *Previous hospitalization in the last 6 months, no. (%)* |  |  |
| 0 hospitalization | 33,341 (66.1%) | 8,543 (65.9%) |
| 1 hospitalization | 8,869 (17.8%) | 2,362 (18.2%) |
| 2+ hospitalizations | 8,157 (16.2%) | 2,060 (15.9%) |
| *Patient type, no. (%)* |  |  |
| Inpatient | 31,875 (63.2%) | 8,179 (63.1%) |
| Observation | 11,926 (23.6%) | 3,120 (24.1%) |
| Same day admission | 6,666 (13.2%) | 1,666 (12.8%) |
| *Discharge destination, no. (%)* |  |  |
| Expired (Death) | 1,113 (2.2%) | 290 (2.2%) |
| Extended care facility | 9,001 (17.8%) | 2,283 (17.6%) |
| Home | 22,672 (44.9%) | 5,780 (44.6%) |
| Home with services | 12,590 (24.9%) | 3,382 (26.1%) |
| Missing | 5,091 (10.1%) | 1,230 (9.5%) |
| *Primary outcomes* |  |  |
| LOS, median (IQR), days | 3.12 (4.42) | 3.12 (4.54) |
| Overall LOS $\geq 14$ days, no. (%) | 3,209 (6.4%) | 844 (6.5%) |
| At least 1 night in ICU, no. (%) | 8,092 (16.0%) | 2,025 (15.6%) |

### 2.2 Modeling and patient representation

Previous approaches (Rajkomar et al. 2018) considered a patient-centric time scale. For each patient, the clock starts at admission and a prediction can be made in the following 12 or 24 hours. On the contrary, we adopt a hospital-centric time scale, where predictions are made on a fixed schedule (daily at 11:59pm in our study) for all inpatients. In our view, this fixed schedule mimics the reality of inpatient management and better captures the periodicity of hospital operations. We define an observation as the state of a patient at the end of each day. For instance, if a patient stays for 5 days at the hospital, she generates 5 observations: one on admission day, one on day 2, and so on. Consequently, the final dataset contains 323, 274 unique observations.

Based on previous work in the literature and discussions with medical staff, we define a rich set of 600+ covariates to describe each observation. These covariates capture the specific condition of the patient (e.g., diagnosis, medications, lab results), her general health state (e.g., BMI, type of diet, level of activity and autonomy) and socio-economic factors (e.g., insurance type, marital status). Most variables are counting operations (e.g., number of blood bank orders submitted in the past day or since admission). For lab results and vitals, we compute daily average, amplitude and trend. We provide an exhaustive list of the data sources and features in Appendix A.

From a scalability perspective, we implement this patient representation natively in SQL so as to easily integrate it into the database infrastructure of the hospital. We are currently working with a second hospital to replicate our approach and assess the degree of similarities between database architectures of different EHR systems. In this regard, we hope standardized format for medical data, such as FHIR (Mandel et al. 2016), will disseminate across institutions and that proprietary systems will become less opaque, for the benefit of the entire industry.

### 2.3 Missing data

Most of the data consists of records of events (e.g., record for surgeries, medical orders, medications) and therefore can not be missing. Some variables, such as homelessness indicator, are mostly missing and equal to “Yes” otherwise, so we consider missing as its own category (“No” in this case). For each patient, lab results and vital signs are imputed using linear interpolation when possible, given their times series nature. Finally, we impute the remaining missing values using an optimization-based imputation method (Bertsimas et al. 2018).

For production deployment, we favor tree-based methods for they can handle missing data by applying conditional mode imputation: At each split, a rule is applied to decide whether to go on the right or left side of the tree. If information required to compute the rule is missing, one decides to go in the direction where the majority of the training observations went. For instance, at Leaf 1 in Figure 2, a prediction is made based on the value of the daily platelet level of the patient. Conditionally on being in that part of the tree, we historically observed 89 patients with platelet level lower than 200.5, and 109 patients with higher platelet levels. Without any information on the platelet level for a particular patient, we will then assume her platelet level is higher than 200.5, as for the majority of past patients.

Note that, apart from regression, all methods in our study are tree-based methods. However, while CART (Breiman et al. 1984) and Optimal Trees produce (Bertsimas and Dunn 2017) a single decision tree, gradient boosted trees (Friedman 2001) and random forest (Breiman 2001) combine predictions from multiple trees, hence losing in interpretability.

### 2.4 Relevant outcomes

We answer four length of stay-related questions. For resource planning purposes, predicting same-day or next-day discharges provides visibility on future bed availability. Accordingly, we build models to predict whether the remaining length of stay was less than 1 or 2 days. From a clinical perspective, it is also useful to identify long-stay patients, i.e., patients whose overall length of stay exceeds 7 or 14 days. Since early detection of long-stay patients can be crucial, we build classification models using only the first four days of each admission to answer those two questions. In other words, if a patient stayed more than four days in total, only the four observations corresponding to the first four days of the admission are kept in the training data. As a result, although the predictive models can be applied at any stage during hospitalization, they are specifically tailored for the beginning of the stay.

We then investigate discharge destination prediction, formulated as a four-class classification problem (between Home, Home with services, Extended Care Facility, Death), with a focus on hospital mortality and extended care facility specifically.

Finally, we predict whether a patient will need an ICU bed in the next 24 hours.

We would like to highlight the fact that some response variables are subjective and may be endogenously determined by physicians. Length of stay, in particular, is first and foremost a result of decisions made by physicians and inefficiencies at the hospital. As a result, machine learning models should not be used as a recommendation engine. At best, they can learn how long a patient will stay, accounting for common protocols and current inefficiencies, not how long she *should* stay. Yet, predicting these quantities, as subjective as they may be, is valuable to hospital managers. Indeed, predictive tools provide estimates on the status of each patient that can then be used as a coordination tool between units, between physicians and other services at the hospital. In complex systems like hospitals, lack of information sharing and irregularities in information reporting are responsible for many of the current inefficiencies, which technologies like machine learning can address.

### 2.5 Model evaluation and statistical analysis

Patients are split based on their admission date into train (Jan - Dec 2017, 60%), validation (Jan - Apr 2018, 20%) and test (May - Aug 2018, 20%) sets. Our evaluation process thus mirrors real-world implementation, where a machine learning model is trained on past data and later applied to future patients. Consequently, we believe our estimates of out-of-sample performance are more realistic than with random training, validation, and test sets. In addition, the common practice of splitting the data at random produces three data sets that are statistically similar, hence leading to overly optimistic performance assessment (see Riley 2019, and references therein). This distinction ought to be kept in mind when comparing our performance with other approaches. Models are trained on the train and validation sets, using the validation set to calibrate hyper parameters and avoid overfitting (holdout method). We provide some background information and implementation details about the machine learning algorithms used and their corresponding hyper parameters in Appendix B. Observations in the train set are weighted according to their class prevalence to account for unbalanced outcomes. We report accuracy metrics computed on the test set only. We assess performance of classification models by calculating the area under the receiver operating curve (AUC). 1,000 bootstrapped samples were used to calculate 95% confidence intervals.

### 2.6 Computing resources

Data preprocessing is done on our partner hospital database, natively in SQL. Train and test of the predictive models are done in Python 3.5.2 and Julia 1.0.1 on a MacBook Pro with 2.5 GHz Intel Core i7 CPU and 16GB of RAM. In other words, our approach is technologically affordable for the vast majority of medical institutions.

## 3 Predictive accuracy on retrospective data

In this section, we report and comment on the predictive power of five machine learning methods. More detailed results are reported in Appendix C.

### 3.1 Predicting imminent discharges

For operational purposes, identifying imminent discharges helps predict future hospital census and bed availability. As in Van Walraven and Forster (2017), we apply a two-stage procedure: We first train machine learning models to estimate the probability of each patient being discharged and assess their performance in terms of AUC. To predict daily discharge volume, we then adjust these probabilities based on the day of the week and sum them over all patients at the hospital. We detail this two-stage procedure in Appendix D. We also refer to Appendix D for details on how we convert predicted probabilities - of discharge for instance - into binary predictions.

With all methods, we detect same-day discharges with very high accuracy (AUC above 80%), with random forest being the most accurate method in our study (95% CI: 0.839-0.847). As a result, we can predict daily number of discharges with a median absolute error (MAE) of 6.2 beds only (IQR: 2.8 beds - 14.5 beds), corresponding to an out-of-sample *R*^2^ of 0.841. Using a gradient boosted trees, we predict next-day discharges with an AUC of 0.822 (0.819-0.826).

Comparison with similar studies is always difficult because of different patient population and accuracy metrics. Yet they provide interesting reference points. For surgical patients only, Safavi et al. (2019) developed a deep-learning approach which predicted 24-hour discharges with an AUC of 0.840 (± 0.08), which is comparable with the accuracy we reach, yet on a wider inpatient population. Though on a different inpatient population, their work surpassed in accuracy the random forest model from Barnes et al. (2016). Recently, Van Walraven and Forster (2017) use non-parametric models to predict the daily number of hospital discharges with accuracy roughly comparable to ours (median relative error: 1.4%; IQR −5.5% to 7.1%). McCoy et al. (2018) forecasted discharge volume in two hospitals using time-series algorithm with a mean absolute error of 11.5 and 11.7 beds respectively (*R*^2^ = 0.843 and 0.726 respectively).

### 3.2 Anticipating long stays

Long stay patients are typically patients with more complex medical or social conditions and consume a large amount of hospital resources so that identifying them early in their stay could be extremely beneficial. As in Rajkomar et al. (2018) we identify patients with an overall length of stay above 7 days. They reported accuracy 24 hours after admission, which approximately corresponds to prediction after two days at the hospital with our modeling choice. In our two works, the logistic regression model already performs well: we reach an AUC of 0.827 (0.820-0.834) after one day and 0.807 (0.798-0.816) after two days, which was comparable with their adaptation of the logistic model from Liu et al. (2010) (95% CI 0.80 −0.84). Having implemented a similar linear benchmark, we are thus optimistic about the comparability of our results. Gradient boosted trees achieves 0.830 (0.822 −0.837) after one day, 0.820 (0.816-0.825) overall, which is comparable with their deep learning approach, without medical notes.

Similarly, we train models to detect overall length of stay above 14 days, a threshold more relevant to our partner hospital, and reach a 0.826 AUC (0.820-0.833) using logistic regression.

### 3.3 Predicting discharge destination

Discharge destination can sometimes be as important as time-to-discharge itself. As presented in Table 3, we are able to accurately classify patients between the four potential discharge destinations with a weighted AUC around 77% for decision trees and 83% for ensemble methods.

**Table 3:** Summary of the results on predicting length of stay (overall and remaining) for logistic regression (LR), CART decision trees (CART), optimal trees with parallel splits (OT), random forest (RF) and gradient boosted trees (GBT). MAE = Median Absolute Error. MRE = Median Relative Error. Full results are reported on Table 12

|  | LR | CART | OT | RF | GBT |
| --- | --- | --- | --- | --- | --- |
| **Classification: remaining length of stay < 1 day** |  |  |  |  |  |
| AUC | 0.826 | 0.807 | 0.810 | **0.843** | 0.839 |
| MAE in # daily discharges, no. | 8.6 | **6.0** | 6.4 | 6.2 | 7.8 |
| MRE in # daily discharges, % | 8.7 | 6.0 | 6.5 | **5.8** | 7.6 |
| Out-of-sample $R^2$ | 0.730 | 0.868 | 0.847 | 0.841 | 0.804 |
| **Classification: remaining length of stay < 2 days** |  |  |  |  |  |
| AUC | 0.809 | 0.786 | 0.790 | 0.815 | **0.822** |
| **Classification: overall length of stay < 7 days** |  |  |  |  |  |
| AUC | 0.818 | 0.775 | 0.776 | 0.813 | **0.820** |
| AUC at day 1 | 0.827 | 0.795 | 0.797 | 0.828 | **0.830** |
| AUC at day 2 | **0.807** | 0.752 | 0.752 | 0.800 | 0.804 |
| **Classification: overall length of stay < 14 days** |  |  |  |  |  |
| AUC | **0.826** | 0.777 | 0.777 | 0.820 | 0.794 |

**Table 4:** Summary of the results on predicting discharge destination for logistic regression (LR), CART decision trees (CART), optimal trees with parallel splits (OT), random forest (RF) and gradient boosted trees (GBT). Full results are reported on Table 13

|  | LR | CART | OT | RF | GBT |
| --- | --- | --- | --- | --- | --- |
| **Discharge Destination** |  |  |  |  |  |
| Weighted AUC | 0.561 | 0.781 | 0.769 | **0.837** | **0.837** |
| **Mortality (with DNR indicator)** |  |  |  |  |  |
| AUC | 0.934 | 0.946 | 0.940 | **0.962** | 0.951 |
| **Mortality (without DNR indicator)** |  |  |  |  |  |
| AUC | 0.855 | 0.836 | 0.838 | **0.905** | 0.848 |
| **Extended Care Facility** |  |  |  |  |  |
| AUC | 0.832 | 0.804 | 0.800 | 0.850 | **0.855** |
| AUC at day 1 | 0.858 | 0.81 | 0.805 | 0.869 | **0.873** |
| AUC at day 2 | 0.846 | 0.803 | 0.806 | 0.861 | **0.868** |

Two destinations require the utmost attention: Extended care facility, because they trigger additional case management effort, and death. We build one-versus-all classifiers for those destinations specifically and reach an AUC of 0.855 (0.852-0.858) and 0.962 (0.959-0.964) respectively.

For extended care facility, the AUC after the first two days is already above 85%, which would enable anticipating administrative bottlenecks early in the stay.

For hospital mortality, our approach outperforms previous studies (Tabak et al. 2014) and even competes with the deep learning model from Rajkomar et al. (2018) (95% CI: 0.92 −0.96). We notice that the most predictive factor was the presence of a Do Not Resuscitate’ document in the patient’s EHR. One might argue that this feature is too related with the output of interest to be included in the model. To this end, we also train models which did not include this variable and still demonstrate around 90% AUC.

### 3.4 Predicting intensive care need

Finally, the vast majority of patient flows between units corresponds to a change in level of care, that is flows into or out of intensive care units. Accordingly, we predict whether a patient will need ICU in the next 24 hours. As reported in Table 5, we reach an overall AUC above 95% with all methods. This high degree of predictability is due to the presence of standardized protocols which make some patient trajectories more deterministic. For instance, cardiac surgical patients are often admitted to an ICU after surgery.

**Table 5:**
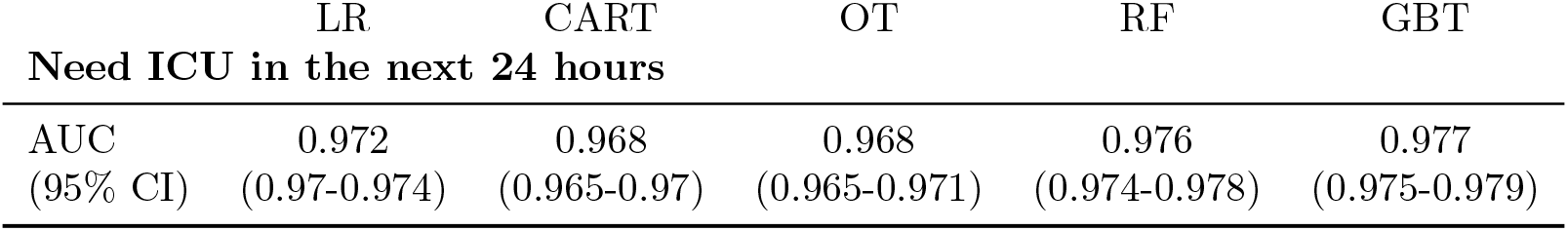
Out-of-sample AUC for predicting ICU need using logistic regression (LR), CART decision trees (CART), optimal trees with parallel splits (OT), random forest (RF) and gradient boosted trees (GBT).

### 3.5 Discussion and limitations

As far as predictive accuracy is concerned, ensemble methods, namely random forest and gradient boosted trees, are the best performing methods. Yet, simple linear regression is already within 4% from the best performing method. Predicting long stays seemed notably harder than predicting imminent discharges, which could be explained by the fact that patients need to reach fairly standardized milestones in order to be discharged while complex cases are a very diverse population.

In our opinion, the value of machine learning is to be found elsewhere than plain accuracy. Linear regression and ensemble methods, like neural networks, can only provide a list of variables ranked by their relative importance. For instance, Table 6 presents the five most important variables for predicting whether the overall length of stay will be above 7 days using a random forest. Table 11 in Appendix C reports a detailed analysis (with coefficient estimates and robust standard errors) of a linear logistic regression model for predicting same-day discharges. In particular, this information is not patient-specific and does not capture non-linear effects. Decision trees, on the other hand, explicit the underlying mechanisms guiding the prediction, while demonstrating satisfactory accuracy. For each patient, a tree provides not only a prediction, but also the path leading to that prediction, which informs both statistical modeling and the care provider: Figure 1 displays the optimal tree predicting hospital mortality with a 0.942 AUC. From this tree, we were able to pinpoint the major predictive power of the Do Not Resuscitate variable and question the use of this variable in our final model. On a similar note, Figure 2 displays particular leaves of the tree predicting long-stay patients, highlighting some clinical (low platelet level, high systolic blood pressure) as well as operational (low discharge volume on Friday-Sunday) barriers to discharge. Of course, those leaves are only valid considering the whole path which leads to them, so conclusions based on those observations should not be drawn too quickly. Interpretability is also a useful tool to avoid bias (Gianfrancesco et al. 2018).

**Figure 1:**
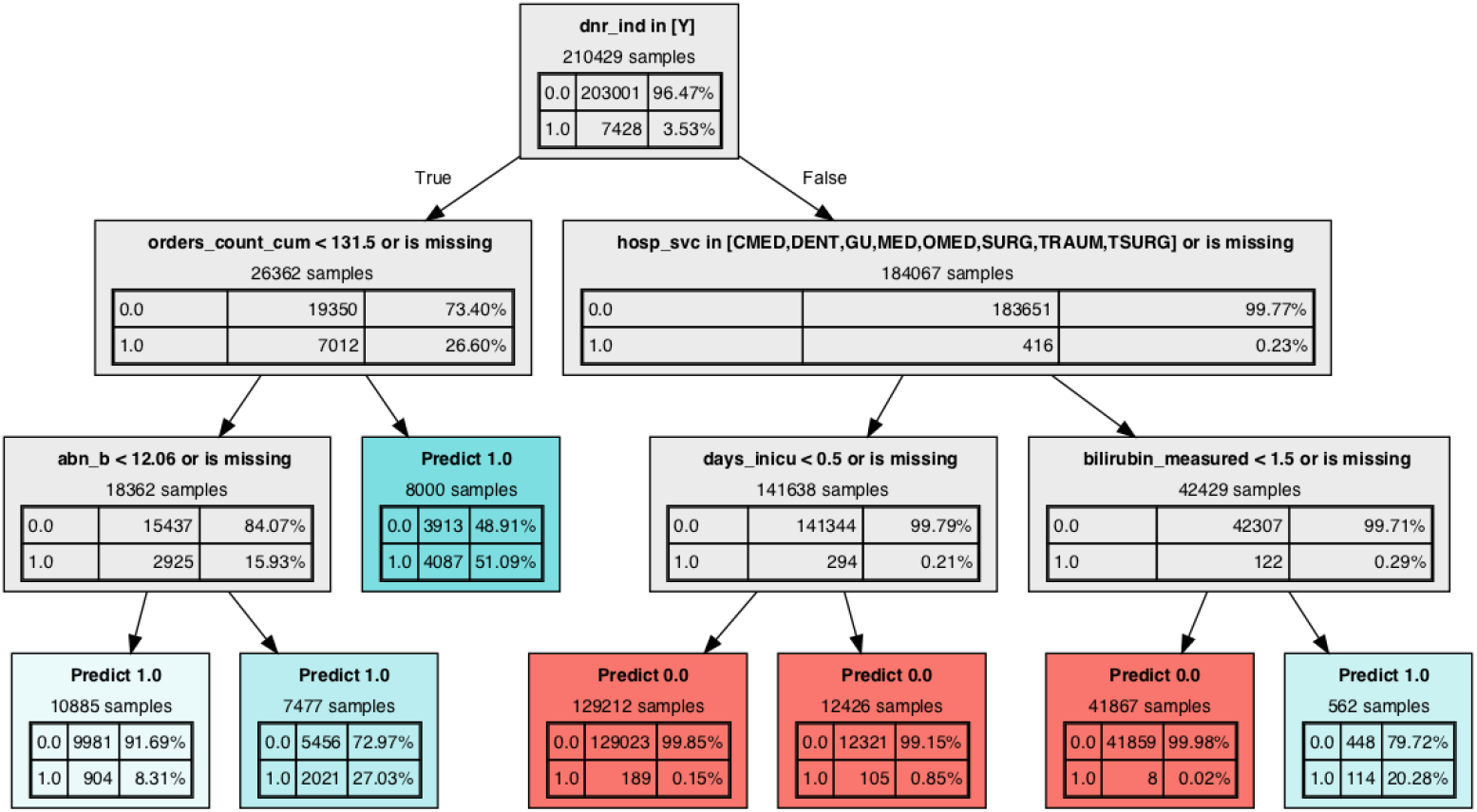
Decision tree predicting inpatient mortalityThe method identifies 6 relevant variables. `dnr_ind:` Indicates whether the patient signed a Do Not Resuscitate form (Y/N) `orders_count_cum:` Number of medical orders placed for the patient since admission. `abn_b:` Number of abnormal blood tests received within the past 24 hours. `hosp_svc:` Hospital service the patient is in. `days_inicu:` Number of days spent in the ICU since admission. `bilirubin_measured:` Indicates whether bilirubin level is measured (0: not measured, 1: as model decalibration and retraining.

**Figure 2:**
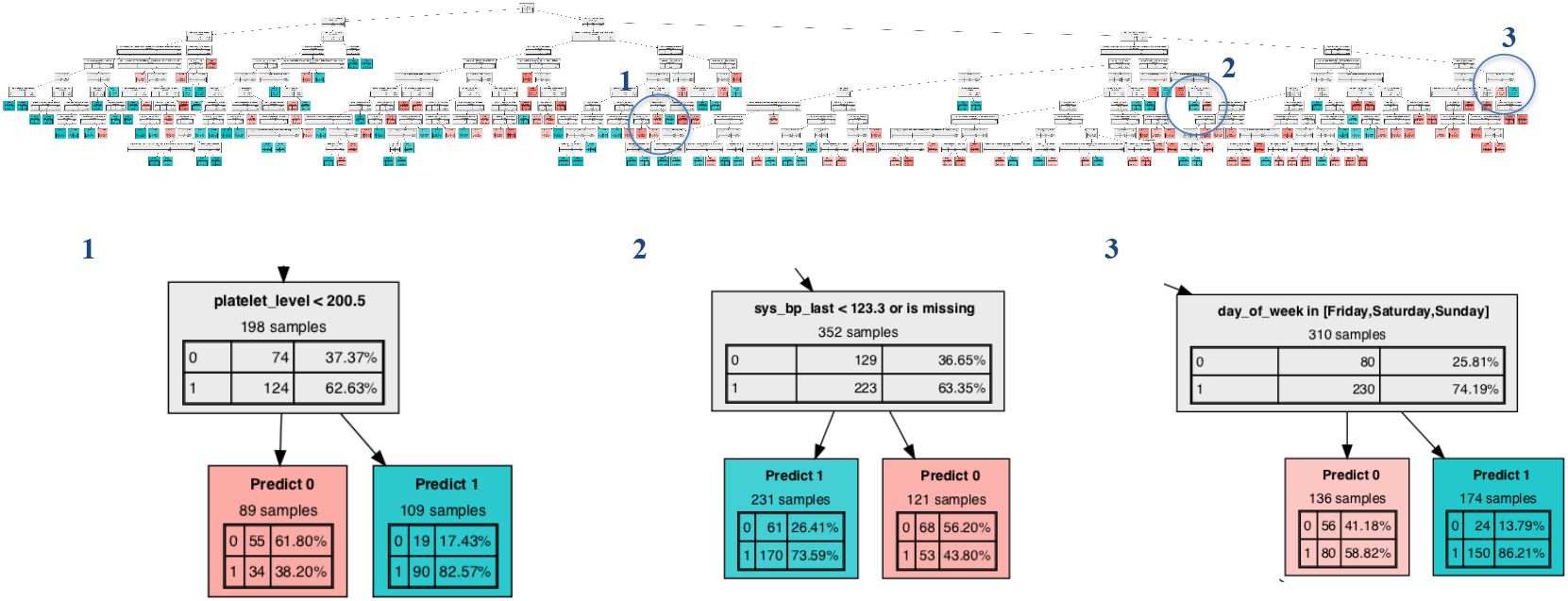
Look-up at a decision tree for predicting whether a patient will stay more than 7 days on overall. Class 0 corresponds to long stays, while class 1 corresponds to lessthan-7-day stays. Leaf 1 suggests that patients with low platelet level (`platelet_level` < 200.5 *k*/*µL*) experience longer stays. Leaf 2 identifies patients with higher-than-normal systolic blood pressure (`sys_bp_last` > 123.4mmHg) as more likely to experience long stays. Leaf 3 confirms that end of the week (when day of the week is Friday, Saturday or Sunday) is less prone to discharges.

**Table 6:** Top 5 variables selected by the random forest classifier for predicting whether a patient will stay more than 7 days on overall.

| Rank | Variable | Description |
| --- | --- | --- |
| 1 | orders_count_cum | Number of orders placed since admission |
| 2 | lab_orders | Number of lab orders placed in the last 24 hours |
| 3 | mars_count | Number of MARS documentation written in the last 24 hours |
| 4 | abn_b | Number of abnormal blood results received in the last 24 hours |
| 5 | orders_types_cum | Number of unique order types placed since admission |

Our study has several limitations. For now, our analysis is single-center only and we are working with a second hospital to adapt and validate the benefits of our approach. Second, any statistical model is only as good as the data it is trained on. Consequently, when predicting time-to-discharge, our models incorporate operational inefficiencies. In presence of undesirable readmissions or administrative delays, it is unclear whether length of stay alone is a relevant outcome to consider. Yet, from an operation standpoint, length of stay remains a key metric and interpretability of the models allow clinicians to read predictions with a pinch of salt. In addition, our approach brings a valuable piece to the overall length of stay vs. readmission risk trade-off. Thirdly, operational deployment of machine learning methods on EHR data requires advanced electronical data collection and management, and reduces the scope of usable information, such as medical notes.

## 4 Deployment in production

In this section, we present how we implement our machine learning pipeline into the EHR system of the hospital and integrate our predictions in daily decision-making. We also report out-of-sample performance in production and discuss practical implementation issues such

### 4.1 Implementation bottlenecks

Besides predictive power, our work demonstrates the value of careful modeling and interpretability. Rajkomar et al. (2018) proposed a generic data processing pipeline to alleviate the burden of data cleaning and modeling, which they consider as the major implementation bottleneck. Our experience has been different.

Their pipeline, though viable for a retrospective study, requires immoderate computing power and daily available medical notes. We show that, even without text notes, a tailored modeling can incorporate clinical expertise from physicians, nurses and case managers to reach comparable performance and increased engagement. Since our model updates predictions on a fixed schedule (every day at 11:59 pm), it can easily be synchronized with operations on the floors (shifts, doctor rounds) as well as with the IT schedule for data backups and updates.

Furthermore, as opposed to tree-based methods, deep learning approaches are not interpretable by design. Rajkomar et al. (2018) use attribution methods to highlight the elements from a patient’s EHR which impacted prediction. This visualization procedure required retraining a model on a restricted version of the data, so the depicted model differs from the one making the predictions. On the contrary, decision trees are easy to understand, even by medical staff who are not trained in machine learning. Actually, the hospital only implemented models based on a single decision tree (CART or OT)^1^for they did not view the gain in accuracy worth the loss in interpretability. However, single-decision-tree methods are notoriously unstable to perturbations in the data, due to the discrete nature of the splits. Such instability can seriously question their interpretability appeal in practice. To assess robustness and stability of the trees produced, we follow the approach of Zimmermann (2008). Namely, we conduct a 10-fold cross validation and compare the depth of the trees obtained on each fold. A method can be called stable if the best depth selected on each fold independently remains constant. As depicted on Figure 3, as the size of the data increases, the method stabilizes: namely the average best depth selected on each fold stabilizes around 5.5 with 6 months of data and more, and the dispersion around the mean shrinks and decreases from 3.7 with 3 months of data down to 0.7 with 6+ months. Consequently, with 16 months of data in the training and validation sets, we are confident in the robustness of the trees obtained in this manner. Yet, we acknowledge the fact that other hyper parameters or semantic stability could be used to quantify stability of decision trees (see Mirzamomen and Kangavari 2017, and references therein). Trading-off stability and interpretability of decision trees remains an active and practically relevant area of research (Last et al. 2002, Mirzamomen and Kangavari 2017, Wang et al. 2018).

**Figure 3:**
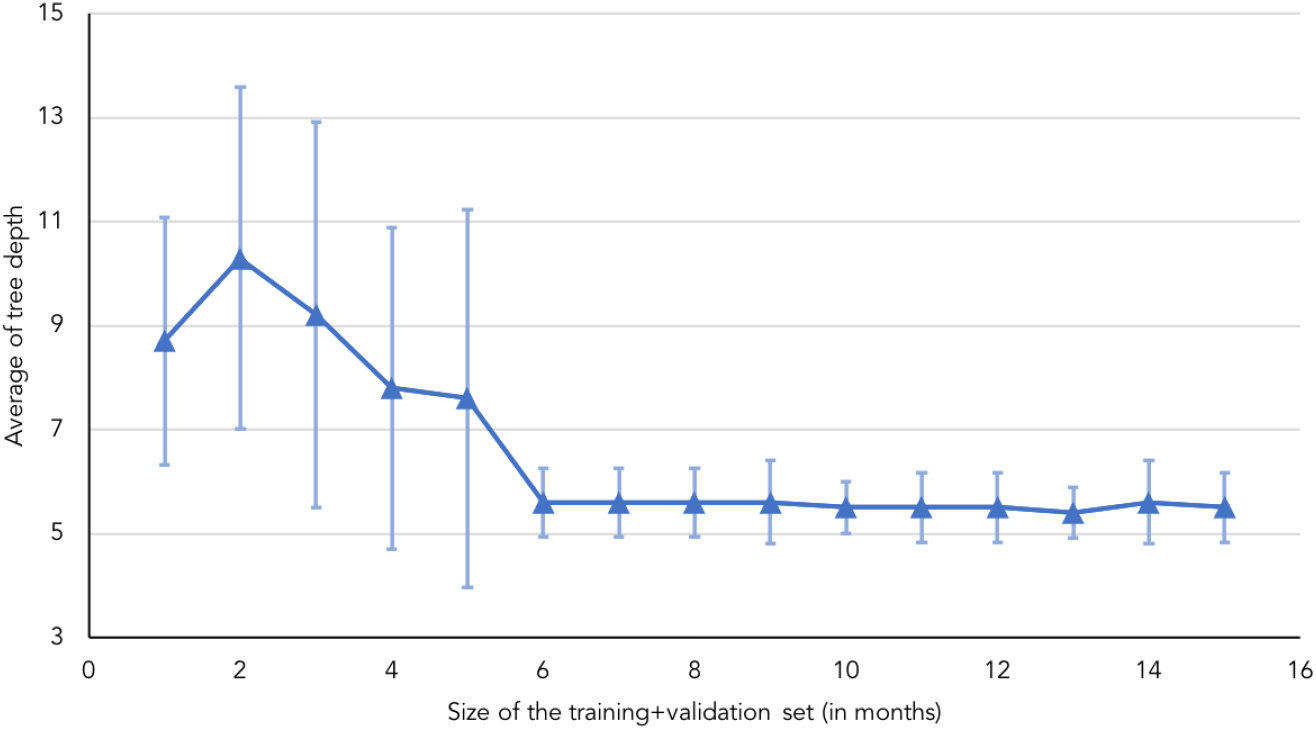
Average depth of the tree selected over 10 random training/validation splits of the data, as the size of the data set (in month) increases. Error bars correspond to ± 1 standard deviation. The prediction task considered is next day discharges, *rLOS* < 1 day.

### 4.2 Machine learning-enhanced dashboards

Once machine learning models are deployed in production in the EHR system, we build interactive dashboards to communicate the output of the model to practitioners.

We work with the office of bed management and design a dashboard where all the hospital wards are listed alongside the census level and estimates for daily discharges and ICU patients (Figure 4). This information is available every morning at 7 am. This is a critical time for patient flow management because bed requests from the emergency department accumulate during the night and most of the discharges will happen only later in the day. With this decision-support tool, practitioners have visibility on the discharge level to be expected for the day and make informed bed assignment decisions. Figure 5 compares the predicted and observed discharge volume in the Summer 2018 (end of the test set) and illustrates how accurately our tool can be used to anticipate daily discharge patterns.

**Figure 4:**
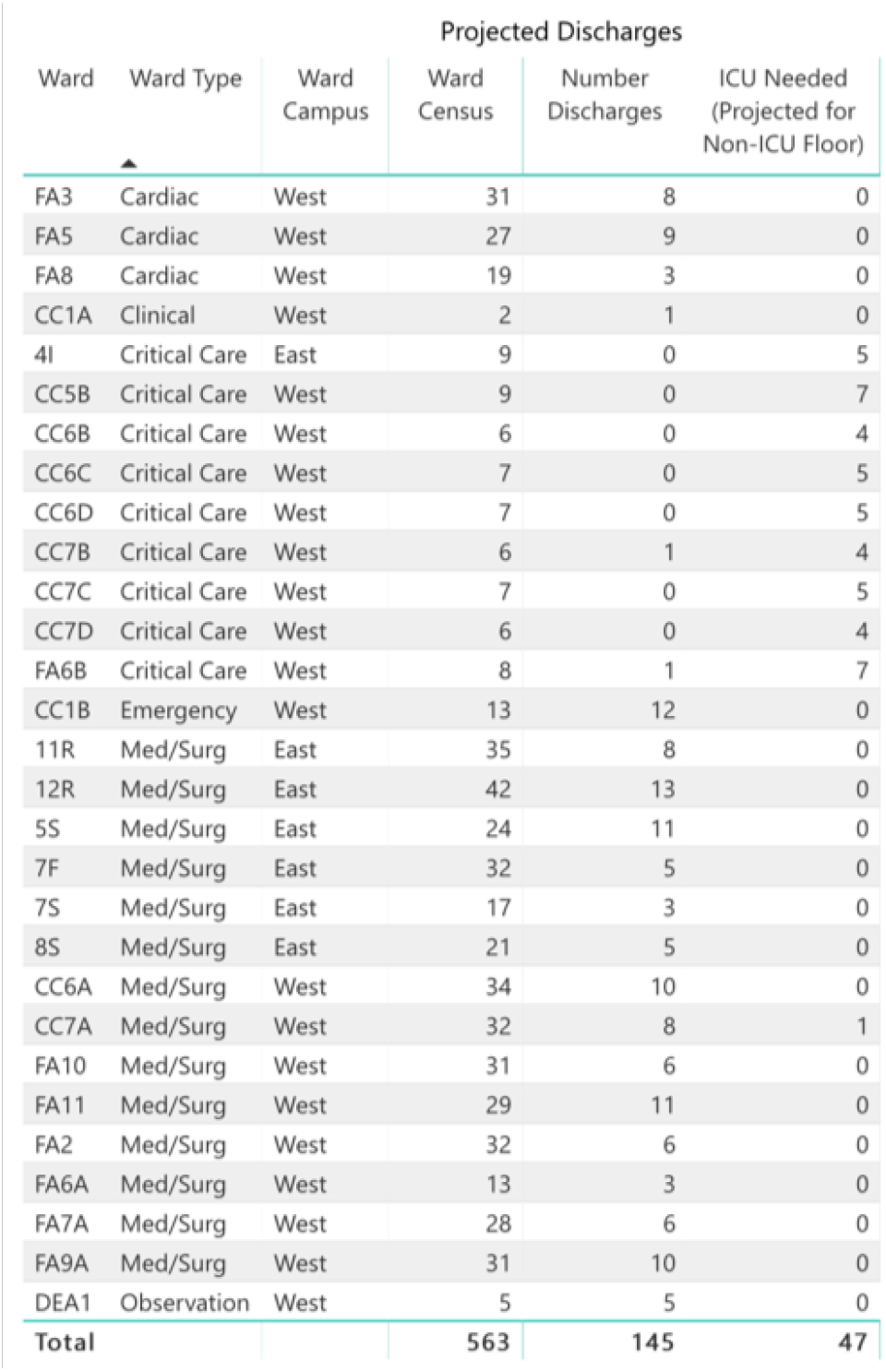
Screenshot of the capacity prediction tool built for the office of bed management. The dashboard displays a list of all the hospital wards with census level, expected number of discharges and expected number of ICU patients by the end of the day.

**Figure 5:**
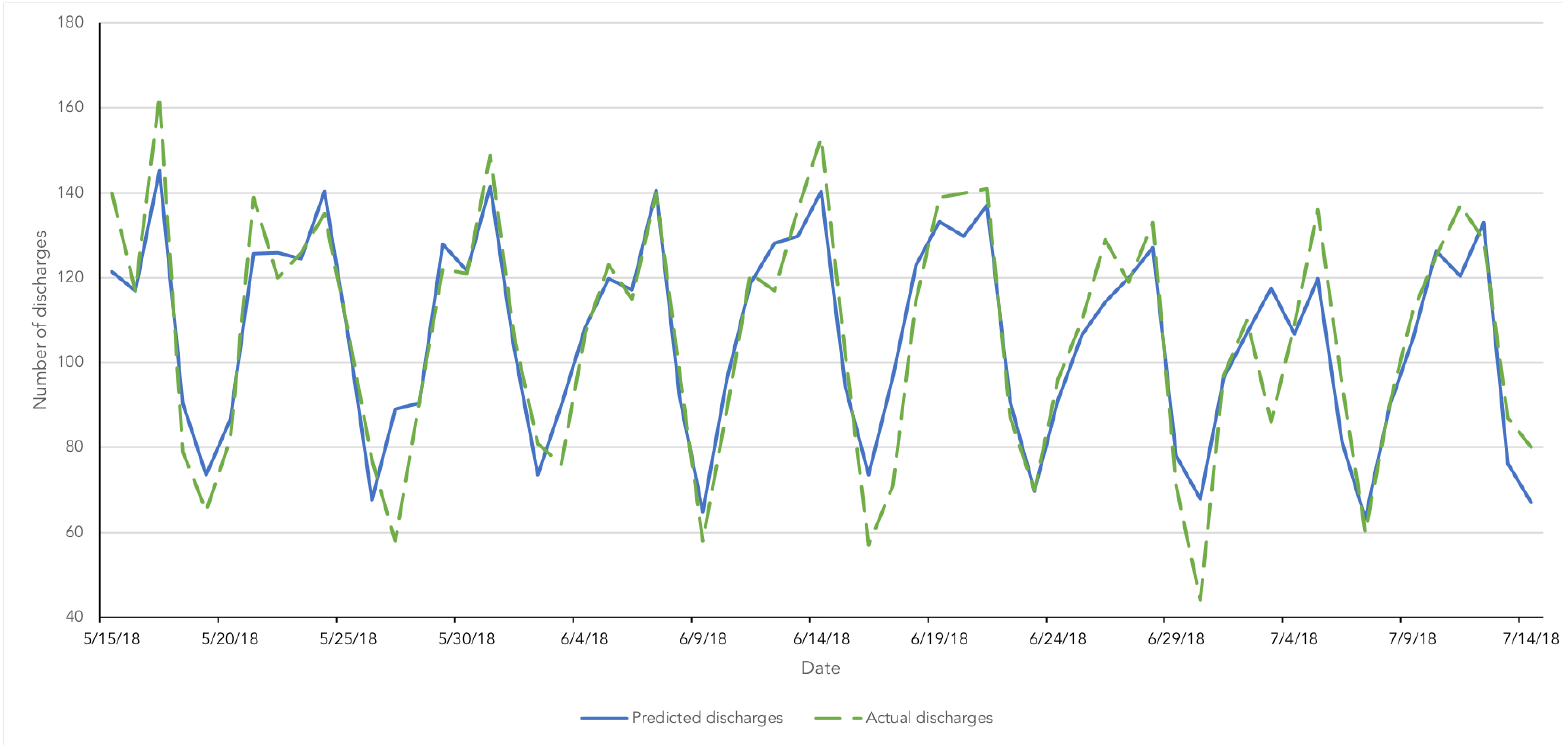
Comparison of actual and predicted discharge volume between mid-May 2018 and mid-July 2018.

### 4.3 Operational impact

In this section, we design and conduct an experiment to assess the operational impact of the tools we develop as rigorously as possible. Since our models are predictive only, we are not making any precise recommendation and do not translate predictions into precise guidelines on how to improve patient flows. Instead, we are relying on the assumption that practitioners will make better informed, hence better decisions. We confront this assumption here with empirical evidence from our partner hospital and its operations with and without our tool.

We deploy the decision-support tool in May-June 2019. On 15 out of those 42 days (weekends and holidays excluded), the nurse in charge of the office of bed management used the tool and reported its prediction, alongside estimates obtained from individually asking resource nurses for predictions on their respective floor. For these 15 days, we can compare the accuracy of our model with the estimates obtained from the resource nurses, on predicting overall discharge volume. Results are presented in Table 7. Machine learning models provide estimates that are significantly more accurate than resource nurses (median relative error of 11.5 vs. 16%). However, the error level achieved by optimal classification trees (11.5%) are noticeably higher than the levels reported in our retrospective study in Section 3, Table 3 (6%). This under-performance is due to decalibration of the model over time and can be fixed by retraining the model regularly on more recent data. We investigate this procedure further in the next section. In addition to accuracy, machine learning models are easier and more consistent than resource nurses. Indeed, collecting estimates from nurses require time to manually inquire each of them, an effort that bed management officers usually do not have time to do. In addition, nurses report predicted discharge volume as intervals^2^whose width greatly vary based on their individual experience. On that regard, practitioners value quantitative models for their predictability and constant behavior.

**Table 7:** Predictive error of the machine learning models vs. resource nurses on hospital discharge volume, during the treated days. Error is measured in terms of relative error compared with actual discharge volume

| Method | 1st quartile | Median | 3rd quartile |
| --- | --- | --- | --- |
| Resource nurses | 5.3% | 16.0% | 20.3% |
| ML model (OCT) | 8.9% | 11.5% | 13.3% |

Regarding operational metrics, we analyze boarding of patients from the Emergency Department (ED) to the main hospital. We focus on the management of ED patients for their are the priority of the office of bed management, who is in charge of placing these patients into inpatients units and who constitutes our targeted end-user. In particular, we are interested in improvements in terms of boarding delays, i.e., the time a patient waits between request for an inpatient bed and admission to an inpatient ward, and off-service placement, i.e., whether the patient is admitted to a unit corresponding to the medical specialty she needs.

To assess the impact of our approach, we perform a difference-in-differences analysis (see Lechner et al. 2011, for a review): We compare daily boarding of ED patients in April 2019 (before the introduction of the dashboard) with July 2019 (after intervention). However, difference in these metrics between April and July 2019 could be due to monthly seasonality. To account for this limitation, we use 2018 data as the control group. More precisely, we assume that the difference in behavior between April and July would have been similar in 2018 and 2019 without the introduction of our solution (parallel trends assumption). In our regression, we also control for day-of-the-week and hour of the day. We summarize the output of our difference-in-differences analysis in Table 8. Notably, we observe that our intervention reduces the proportion of off-service placement by 4% (p-value 0.0095 < 0.01).

**Table 8:** Difference-in-differences analysis of boarding delays and off-service placement between April and July 2019. We use April and July 2018 as control data. The variable “2019 indicator” captures the changes in activity between 2018 and 2019, “July indicator” captures the monthly seasonality between April and July, and “2019 indicator × July indicator” captures the effect of our intervention. We report estimates for each coefficient (with robust standard errors) and level of significance.

|  | Boarding delay |  | Off-service placement |  |
| --- | --- | --- | --- | --- |
| | Coefficient | $p$-value | Coefficient | $p$-value |
| 2019 indicator | 0.231 (0.122) | $< 0.1$ | 0.053 (0.012) | $< 0.001$ |
| July indicator | 0.442 (0.115) | $< 0.001$ | 0.032 (0.011) | $< 0.01$ |
| 2019 indicator × July indicator | -0.120 (0.174) | | -0.043 (0.017) | $< 0.01$ |
| Controls: day-of-the-week, hour of the day |  |  |  |  |
| Observations: 5,126 |  |  |  |  |

To affranchise our result from the parallel trends assumption, we also conduct a more granular analysis during the implementation period, i.e., May-June 2019. As previously mentioned, nurses at the office of bed management reportedly used the dashboard on 15 out of 42 days. We refer to these days as the “treatment group”. Assignment to treatment was mostly driven by system development and maintenance to improve the dashboard, and can therefore be considered as independent from hospital’s activity level. However, we acknowledge the fact that nurses might have used the dashboard without reporting (leading to mislabeling between control and treatment group) and that failure to use/report can be correlated with hospital’s activity (leading to correlation between treatment assignment and outcome of interest). We verify that treated and control days are reasonably comparable: Median number of ED visits in the treated and control group is 159 (IQR: 146.5-167.0) and 154.5 (IQR: 148.2-163.5) respectively; median census is 712 (IQR: 699-721) and 698.5 (IQR: 688.5-717) respectively.

We analyze inpatient admissions from the ED during this period. To account for seasonality in patients arrivals and hospital operations, we stratify these 2,117 patients based on the time when they request for a bed. Specifically, we construct 120 groups based on the day of the week and hour of the day and exclude 4 groups which contained control patients only. We compute average response to treatment/control within each group and report summary statistics of the average treatment effect across strata in Table 9. We consider two outcomes: boarding delay and off-service placement. For these two metrics, we observe an improvement in the majority of cases (nonpositive median treatment effect). These observations remain valid if we consider as “treated” a day where the tool was used and the prediction was reasonably accurate (within 16% of the truth). Such empirical evidence supports our working assumption: practitioners are most of the time able to leverage more accurate estimates on hospital discharges to improve patient management. Yet, implementing a predictive analytics tool in a real-world medical environment over which we have little control makes rigorous empirical investigation challenging. In particular, assignment to treatment is ultimately up to nurses on the floor and inherently noisy. Treatment assignment and the limited number of patients constitute, in our opinion, the main limitation of this empirical validation, compared with the difference-in-differences analysis. Further work in a more controlled setting could provide a more robust estimate of the benefit from our tool.

**Table 9:** Operational impact of the adoption of our machine learning-based dashboard on patient admission in May-June 2019. We compute Average Treatment Effect (ATE) over 116 strata, based on day of the week and hour of the day, for two operational metrics.

| Response | Median ATE [IQR] | $\mathbb{P}(ATE < 0)$ | $\mathbb{P}(ATE > 0)$ | $\mathbb{E}[ATE\vert ATE < 0]$ | $\mathbb{E}[ATE\vert ATE > 0]$ |
| --- | --- | --- | --- | --- | --- |
| Boarding delay (in hours) | -0.18 [-2.02 ; 1.21] | 55% | 45% | -2.26 | 2.36 |
| Off-service placements | 0.00 [-0.10; 0.11] | 38% | 39% | -0.15 | 0.18 |

Furthermore, we believe that predictive tools could be built on top of such prediction engines to convert our predictive tool into actionable recommendations, improve patient management further, and lead to even greater impact.

### 4.4 Model maintenance

Now that machine learning models are deployed in production and inform decision-support tools, natural questions arise at an organization level on how to maintain and update such models over time. From an applied machine learning perspective, two questions need to be answered: how often should the model be retrained? And how does the amount of training data impact predictive accuracy?

Regarding the first question, we have run the models in production for a year and report the relative error in daily discharges over time (Figure 6). Empirically, we observe that (a) the model does not capture well exceptional events such as holidays, as demonstrated by the high error level around the end-of-the-year holiday season for instance, (b) the relative error increases roughly by +1 percentage point every three months (b) the error levels are in par with out-of-sample performance on the test set until December 2018, i.e., 8 months after the end of the training + validation set.

**Figure 6:**
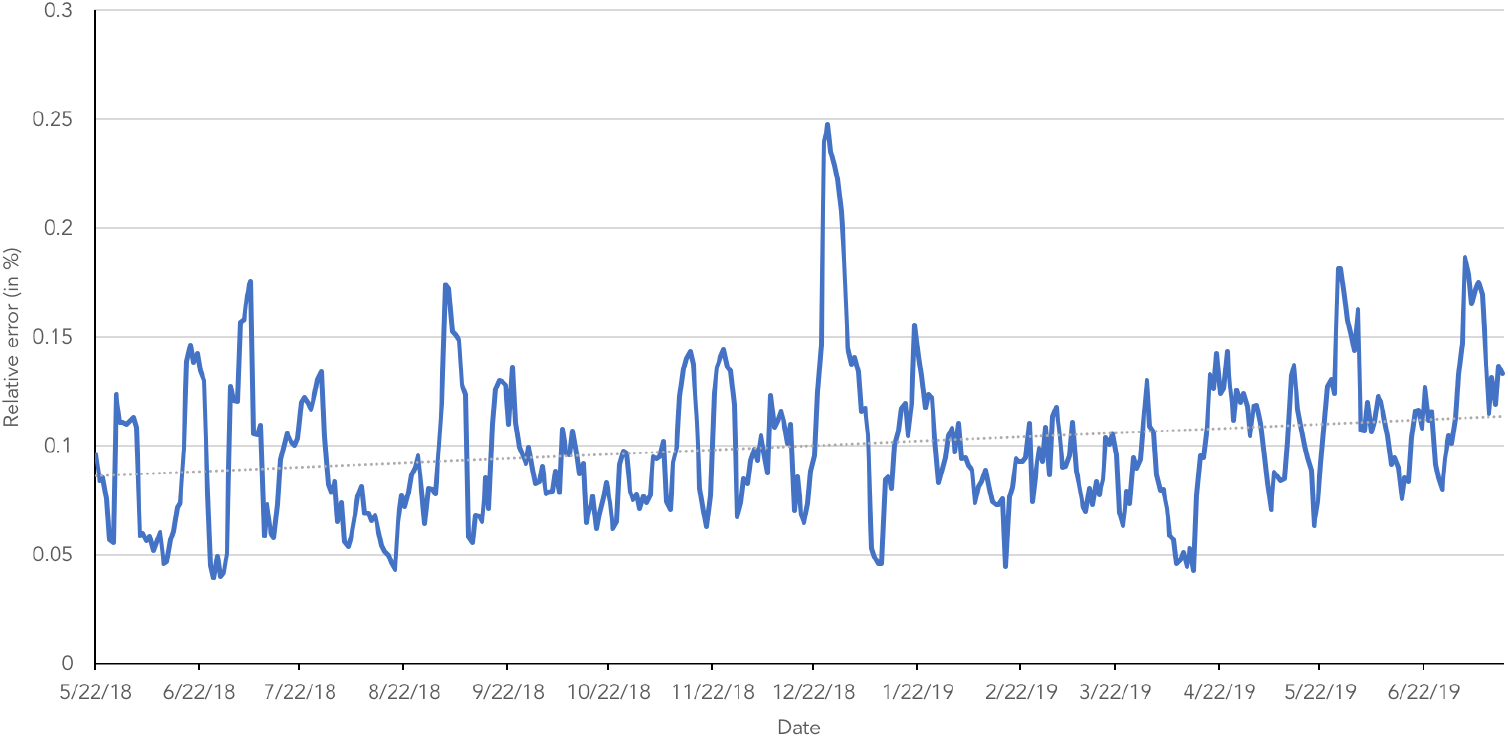
Running average (7-day window) of the absolute error in discharge prediction over a year.

To assess the “marginal value of data”, that is how increasing the size of the training set impacts prediction accuracy, we adopt the following methodology: We consider the patients admitted between January and August 2019 as a test set and we train machine learning models from a training and validation set consisting of *m* contiguous months ending in December 2018, where *m* varied from 1 to 24 (2 years). We report out-of-sample AUC for a CART decision tree and a random forest model as *m* increases for same-day discharges prediction on Figure 7. As expected, predictive power is an increasing and concave function of the training period size. For this particular task, it seems that around 14 months of data is needed to reach full accuracy, which luckily coincides with the size of our training + validation set for our original retrospective study (16 months).

**Figure 7:**
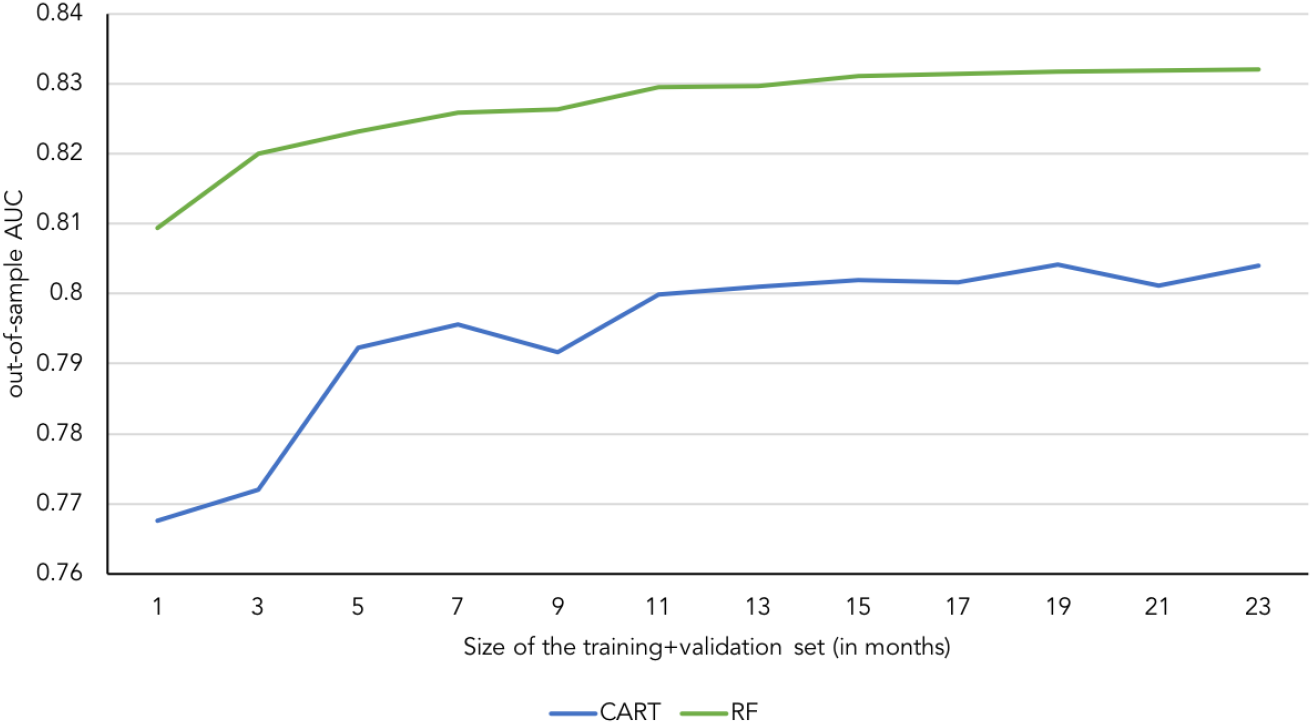
Running average (7-day window) of the absolute error in discharge prediction over a year.

Based on these observations, we advise to our partner hospital to retrain the models biannually, updating the training data with observations from the past semester while keeping the training period size roughly constant. Naturally, these guidelines are problem-and institution-specific but these ex-post performance analyses constitute best practices that we passed on to our partner organization and that are required for any sustainable and effective analytics strategy.

## 5 Conclusion

In this study, we demonstrate how tailored modeling can be used in combination with interpretable machine learning techniques to provide accurate predictions on critical aspects of patient flows. To the best of our knowledge, our study is first of its kind to (a) address the length of stay and discharge destination prediction task for such a generic inpatient population with a unified data modeling and processing, (b) achieve state-of-the-art accuracy with a broad collection of models, including interpretable ones, (c) be fully integrated into the EHR system of a major hospital, thus demonstrating how powerful analytics can concretely impact care delivery. Yet, further work is needed to ensure and validate that accurate predictions translate into improved quality of care.

## Data Availability

Data is not available

## A Construction of the patient representation

In this supplement, we describe the features we created from the raw EHR data to model the state of each patient.

We divide the variables into two categories:

1. static variables, which are not supposed to change as the patient stays at the hospital,
2. dynamic variables, which are daily updated.

### A.1 Static variables

Static variables represent a patient’s socio-economic situation, as well as existing conditions. We assume these variables do not evolve over a patient’s stay. We acknowledge that this assumption might not always hold in practice. For instance, for certain conditions, weight can drastically change over one’s stay. In addition, some data such as insurance information might not be available and entered in the EHR system directly at admission.

Static variables are obtained from three sources, namely

- Admission data,
- ICD10 codes from previous admissions (billing data),
- Initial Patient Assessment information (completed on the day of admission), and consist of
- Admission data

– Patient’s type and source
– Hospital service responsible for the patient
– Age
– Gender
– Weight, height, BMI
– Insurance indicators
– Homelessness indicator
– Income category based on ZIP code
– English-speaking indicator
- ICD10 codes from previous admissions (billing data)

– Indicator for each comorbidity, based on Elixhauser comorbidity software https://www.hcup-us.ahrq.gov/toolssoftware/comorbidity/comorbidity.jsp
– Indicator of chronic condition, by body system, based on the CCI classification https://www.hcup-us.ahrq.gov/toolssoftware/chronic_icd10/chronic_icd10.jsp
- Initial Patient Assessment information (completed on the day of admission)

– Living situation (alone, family, group setting)
– Indicators of autonomy level (ability to use bell, autonomy in activities of daily life, difficulty ambulating or swallowing)

## A.2 Dynamic variables

The dynamic variables we consider are:

- From general admission data

– Number of days the patient already spent at the hospital during this admission,
– Daily number of admission/discharges at a hospital/ward level,
– Daily number of (long-stay) patients at a hospital/ward level,
– Current day of the week,
– Current service the patient is in. This variable is used to identify off-service patients,
– Indicator of whether the patient is currently in an ICU,
– Number of days spent in the ICU so far,
– Number of days spent with the same attending,
- From vital sign measurements

– We monitor heartrate, respiratory rate, temperature, O2 saturation, diastolic/systolic blood pressure, pain level, activity level, RASS score.
– For those vital signs, we report last value at the end of the day, average daily value and trend based on past 4 measurements.
- From lab result information

– We only include blood results for they are widely performed for all inpatients.
– We count the number of lab measurements at an abnormal value.
– For platelet count, white cells count, hematocrit, chloride, sodium, potassium, RDW, RDW SD and urea nitrogen, we report the last value, the last slack compared to the normal range and the slope based on previous measurements.
– For bilirubin, glucose, PTT, sedimentation rate, troponin, albumin and INR, we create a discrete variable taking values 0/1/2: 0 if the quantity has never been measured for the patient; 1 if the quantity has been measured and currently at a normal level; 2 if the quantity has been measured and currently at an abnormal level.
- From medical orders

– We divide orders into types: Blood bank, Cardiology, Consultation, Critical Care, General Care, Lab, Neurology, Nutrition, Obstetrics, Radiology, Respiratory, IV. Observe that each order can be a new order or a change/termination of a previously placed order.
– For all order types, we daily count the number of orders placed (counting +1 for new and −1 for discontinuing orders) and the number of order changes.
– For some types, namely IV, TPN, Blood bank, we perform those counting operations at a sub-type level as well.
- From pharmacy data

– We group medications based on the AHFS Pharmacologic-Therapeutic Classification (first two digits of the AHFS code).
– Within each group, we compute the number of medications the patient was currently taking, the number of medications the patient started/stoped taking on each day and the time (in days) she had been taking these medications for.
- For diagnosis codes

– We use the number of ICD10 codes reported since admission as a measure of a patient’s severity.
– We group ICD10 codes based on their letter and 1) compute the proportion of codes within each category to identify the type of illness, 2) compute the gini score of the distribution of code letters to measure clinical complexity.
- From the OR schedule

– Future surgery: we compute and report the inverse of the time-to-next-surgery. This quantity equals 0 if no surgery is scheduled.
– Past surgeries: we count the surgeries the patient has had since admission (by number and duration). We report the inverse of the time-to-last-surgery as well.
- From documentation and notes

– Without using raw information from the documentations or notes about the patient, we record the number of documents/notes entered about the patient on each day, to measure the level of “activity” around each patient.
– We also compute the inter-day difference to capture variation in activity.

## B Background on machine learning tasks

In this section, we provide some background information about the machine learning techniques used in our analysis. This appendix is intended as a light introduction to the main concepts of machine learning we will refer to during our analysis. For a more detailed introduction, we refer to Chapters 3 and 4 in Bishop (2006) and to Rokach and Maimon (2008).

### B.1 Empirical risk minimization framework

We are in presence of a supervised learning task where we have access to past observations of input data *x_i_* ∈ ℝ*^p^, i* = 1,…,*n* and their associated output *y_i_, i* = 1,…,*n*. In a regression setting, the outputs are continuous, e.g., *y_i_* ∈ ℝ, while in a classification setting, they take discrete values only, e.g., *y_i_* ∈ {±1}. The goal is to predict the output *y* as a function of the input *x*. The “best” predictor *f*(*x*) is computed from data by solving an empirical empirical risk minimization problem

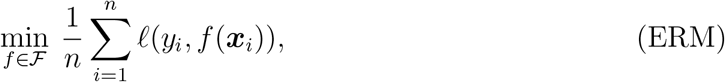

i.e., we seek the function *f*, among a prescribed class of learners 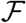, that leads to predictions *f*(*x_i_*) close to the true responses *y_i_*, as captured by the loss function ℓ(·, ·). For regression, the most commonly used loss function is the ordinary least square loss 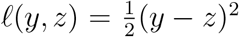. In binary classification settings with *y_i_* ∈ {±1}, taking ℓ(*y*, *z*) = log(1 + *e^−yz^*) leads to logistic regression. For tractability purposes, the optimization problem (ERM) is reduced to a finite-dimensional problem by considering a parametric class of learners 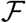 of the form 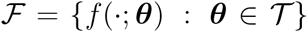, where ***θ*** is a finite-dimensional vector of parameters to calibrate. Hence, minimizing over 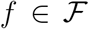 is equivalent to minimizing over 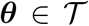. For example, in linear regression, we consider linear functions of the form 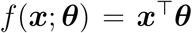, i.e, 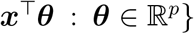 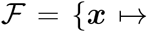.

The outlined approach, however, might lead to over-fitting. Namely, the optimal parameter value for ***θ*** might perfectly explain past observations but perform poorly on newly observed data. To alleviate this issue and achieve higher out-of-sample performance, common practice uses regularization and cross-validation.

Regularization restricts even further the class of learners to 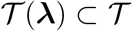, where ***λ*** is a set of so-called hyper parameters that define the set 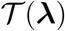. For example in linear regression, Tibshirani (1996) introduced the Lasso estimator for which ***θ*** is constrained in an *ℓ*_1_ sense, namely 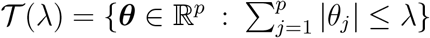.

The standard procedure for finding both ***θ*** and ***λ*** is cross-validation. For a given data set 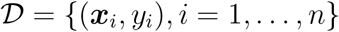 and a parameter ***θ***, define the empirical risk of ***θ*** on 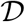 as

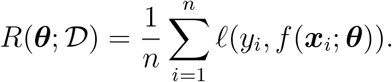

*Hold-out cross-validation* proceeds as follows:

1. Split the original data set into a training and a validation set, 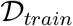 and 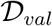.
2. Define a grid of values for ***λ***, {***λ***^(1)^,…, ***λ***^(^*^M^*^)^}.
3. For every value *m* = 1,…,*M*, compute 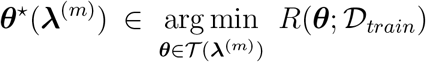, and its out-of-sample performance 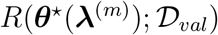.
4. Pick the value of ***λ*** that leads to the best out-of-sample performance 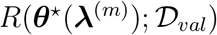.
5. Finally, compute ***θ*** on the entire data set for the best value of ***λ*** = ***λ***^(^*^m^*^)^

**Table 10:** Summary of the hyper parameters involved in each machine learning model and that we tune using cross-validation.

| Method | Hyper parameters |
| --- | --- |
| Linear regression (LR) | Lasso penalty |
| Decision tree (CART and OCT) | Splitting criterion, Maximal depth of the tree, Minimal number of sample per leaf |
| Random forest (RF) | Number of trees in the forest, Maximal depth of the trees |
| Gradient-boosted trees (GBT) | Number of base learners, Maximal depth of the base learners, Learning rate |

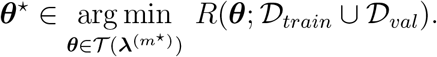

Many variants of this procedure exist. For instance, the loss function used for in-sample and out-of-sample validation can be different depending on the prediction task. Usually, the data is randomly split between training and validation sets, however, deterministic split might not be recommended in cases where the data exhibit known trends (see Riley 2019, for a discussion). When the amount of data is limited, estimation of out-of-sample performance in Step 4 can be improved by splitting the data into training and validation in *k* different ways. Step 3 is performed for each of these *k* splits and out-of-sample performance in Step 4 is averaged over the *k* candidate validation sets. k-fold cross-validation implements such a strategy. In *k*-fold cross-validation, if *k* = n, the validation sets are comprised of a single data point and the method is known as leave-one-out cross-validation.

For our analysis, we perform hold-out cross-validation and split the data according to admission date instead of randomly, so as to mimic real-world implementation and capture non-stationarity if any. We assess out-of-sample performance of classification models on the validation set by the area under the receiver operating curve (AUC), while the loss function for training is the logistic loss.

### B.2 Examples of machine learning techniques

In this section, we review the machine learning techniques we apply in our study. In particular, we emphasize the class of learners 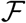 they correspond to, how they are parametrized, and the hyper parameters we tune using cross-validation. We summarize these hyper parameters in Table 10.

#### Linear regression

Linear regression considers linear predictions of the form *f*(*x*; ***θ***)= ***x***^T^***θ***, where ***θ*** ∈ ℝ*^p^*. To avoid over-fitting, we consider the Lasso estimator introduced by Tibshirani (1996). In its original formulation, the Lasso estimator satisfies the constraint 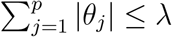 where ***λ*** is the Lasso parameter. Equivalently, the ℓ_1_-norm of ***θ*** can be penalized in the objective of (ERM) instead of being hardly constrained, which is the formulation used in most available implementations.

#### Decision trees (Breiman et al. (1984) and Rokach and Maimon (2008) Chapters 1-7)

A decision tree is defined as a succession of splits similar to the one represented on Figure 8. Each split is defined by a variable *j* ∈{1,…,*p*} to split on and a threshold value *t_j_*. To compute a prediction for a particular data point *x*, we read the first split “*x_j_* < *t_j_*?” and, depending on the answer, go down to the left or right child subtree. We do so until we reach a terminal node, also called a leaf. A prediction is then obtained by taking the average output *y_i_* over all training samples within the same leaf. As a result, decision trees can be parametrized by a finite-dimensional parameter vector ***θ*** that encodes for the feature and threshold of each split. Common hyper parameters that we tune are the maximal depth of the tree (we test all values between 5 and 20) and the minimal number of training samples at each leaf (between 50 and 300). Optimizing over the set of decision trees is a hard combinatorial problem and numerous numerical strategies have been investigated in order to do so, at least approximately. Quinlan (2014) proposed an iterative method to greedily add splits that maximize a given split criterion and then prune the final tree to simplify it and avoid overfitting. Breiman et al. (1984) designed a similar greedy approach where the complexity of the tree is not enforced by pruning but instead penalized in the objective function explicitly (CART). Bertsimas and Dunn (2017) formulated the problem of decision tree learning as a large mixed-integer optimization problem and proposed an efficient local-search heuristic to solve it approximately (OCT). We compare the last two approaches in our numerical experiments for their implementation is easily available, they achieve good predictive accuracy, and do not rely on a pruning step, which generate higher instability in the splits. Yet, due to the discrete nature of the splitting rule, calibration of a decision tree always suffers from instability with respect to the training data (Breiman et al. 1996). This deficiency seriously challenges the interpretability appeal of decision trees in practice and constitues a vivid reserch topic (Last et al. 2002, Zimmermann 2008, Mirzamomen and Kangavari 2017, Wang et al. 2018). All these methods require a criterion to choose the best split, typically the Gini impurity or information gain, which we also consider as an hyper parameter.

**Figure 8:**
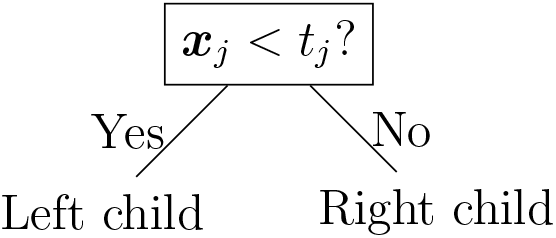
Typical split of a decision tree

#### Random forest (Breiman (2001) and Rokach and Maimon (2008) Chapter 9)

Random forests were initially proposed by Breiman (2001). At a high level, a random forest is simply a collection of trees. Each tree produces a prediction and the final prediction is obtained as the average prediction across all trees in the forest. Using a random selection of features to split each node introduces randomization in the training process of each tree and leads to desirable error rates (Breiman 2001) and consistency results (Scornet et al. 2015). Observe that the training of each tree is independent of the other trees and can be easily perfomed in parallel. Number of trees in the forest constitutes the main hyper parameter to control over-fitting (between 200 and 400 in our grid search). We also tune the maximal depth of each tree (between 20 and 80). We let the other parameters -split criterion, minimal number of samples per split and leaf -to their default value-Gini impurity, 2, and 1 respectively. Each tree in the forest is trained on a bootstrapped sample with replacement of the training data set and of the same size. When looking for the best split within a tree, only *p* features are considered, randomly picked at among the 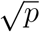 available at each step of the algorithm.

#### Gradient-boosted trees (Friedman 2001)

While random forests rely on averaging a large amount of randomized trees, gradient-boosted trees are an additive method. At each step in the gradient boosting algorithm, a new tree is built so as to predict the residuals, i.e., the fraction of the output that is not explained by the current model. In other words, at each step of the algorithm, a new tree is added to the collection in order to correct for the mistakes made my the current collection. The final output is also a weighted sum of the outputs from each tree. However, while trees in a random forest can be trained independently, trees in gradient boosting can only be trained sequentially. Trees generated at each step are also called base learners and should be of low complexity (we cross-validate their maximal depth between 3 and 7). The total number of trees is another hyper parameter to tune (up to 10). The most important hyper-parameter is the learning rate, between 0 and 1, which controls the contribution of each new tree to the overall model. We let other hyper-parameters-split criterion, minimum number of samples per leaf and split -to their default value -the score proposed by Friedman (2001), 2, and 1 respectively.

## C Extensive comparison of predictive accuracy

In this section, we complement Section 3 by providing a more in-depth assessment of the predictive power of the five machine learning techniques for predicting length of stay-related outcomes (Table 12) and discharge destination (Table 13).

For predicting the number of daily discharges, a regression task answered by aggregating patient-level discharge scores, we report out-of-sample *R*^2^ and adjusted-*R*^2^ in Table 12. The adjusted-*R*^2^, 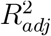 is defined as 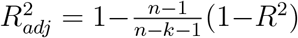, where n is the number of observations in the test set (*n* = 67, 227 here) and *k* is the number of features used by the model (*k* ≤ 700 here). For linear regression, *k* corresponds to the number of covariates whose weights are non-zero. For the other methods, *k* counts the features that appear in at least one split.

In Table 11, we report the detailed output (with coefficient estimates and standard errors) of a linear logistic regression model for predicting same-day discharges (remaining length-of-say < 1 day). In this model, we only included the 50 most important features, as ranked by the random forest classifier. We report the coefficient estimates for the 10 most important ones and use the remaining 40 as controls. As displayed, the regular and robust standard error estimates display no significant difference.

**Table 11:**
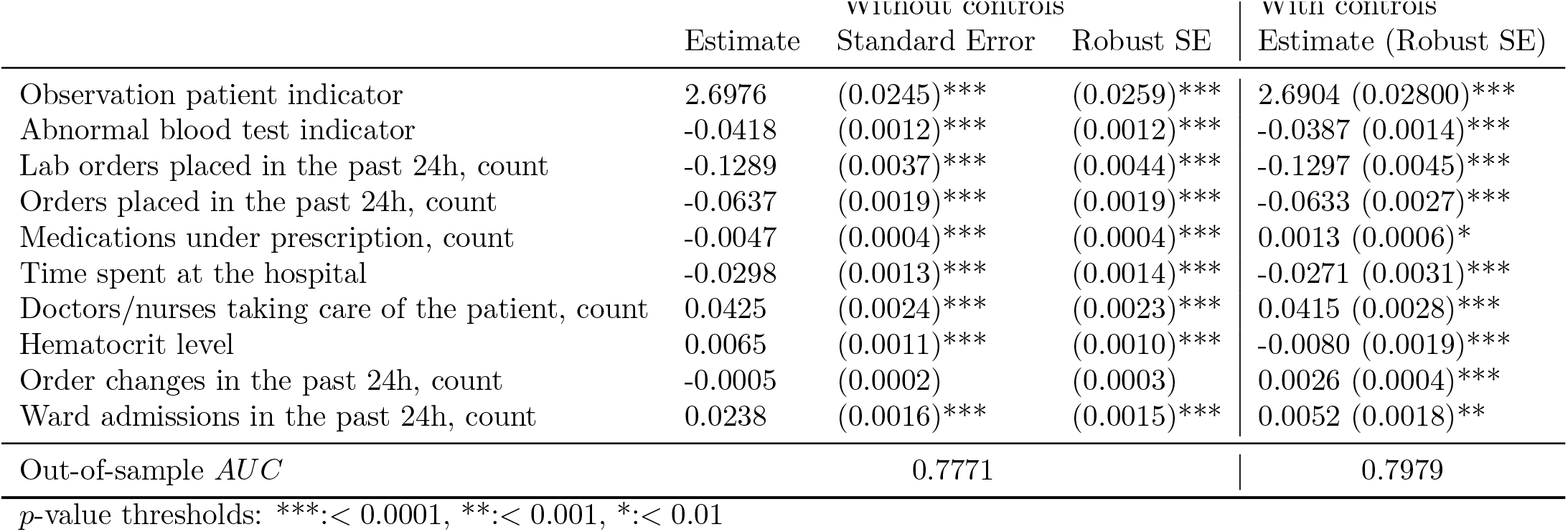
Robust standard error analysis for a linear logistic model to predict whether the remaining length-of-say < 1 day.

**Table 12:** Summary of the results on predicting length of stay (overall and remaining) for logistic regression (LR), CART decision trees (CART), optimal trees with parallel splits (OT), random forest (RF) and gradient boosted trees (GBT). MAE =Median Absolute Error. MRE = Median Relative Error

|  | LR | CART | OT | RF | GBT |
| --- | --- | --- | --- | --- | --- |
| **Classification: remaining length of stay < 1 day** |  |  |  |  |  |
| AUC (95% CI) | 0.826 (0.822-0.830) | 0.807 (0.803-0.811) | 0.810 (0.807-0.815) | **0.843 (0.839-0.847)** | 0.839 (0.835-0.842) |
| MAE in # daily discharges (IQR), no. | 8.6 (4.2- 12.8) | **6.0 (2.5- 11.6)** | 6.4 (3.4- 11.6) | 6.2 (2.8- 14.5) | 7.8 (3.7- 11.5) |
| MRE in # daily discharges (IQR), % | 8.7 (4.1 - 12.9) | 6.0 (2.7 - 12.4) | 6.5 (3.2 - 11.4) | **5.8 (2.3 - 13.4)** | 7.6 (3.3 - 11.7) |
| Out-of-sample $R^2$ / Adjusted-$R^2$ | 0.730 / 0.727 | 0.868 / 0.868 | 0.847 / 0.847 | 0.841 / 0.840 | 0.804 / 0.803 |
| **Classification: remaining length of stay < 2 days** |  |  |  |  |  |
| AUC (95% CI) | 0.809 (0.806-0.813) | 0.786 (0.783-0.790) | 0.790 (0.786-0.794) | 0.815 (0.812-0.819) | **0.822 (0.819-0.826)** |
| **Classification: overall length of stay < 7 days** |  |  |  |  |  |
| AUC (95% CI) | 0.818 (0.714-0.822) | 0.775 (0.770-0.780) | 0.776 (0.772-0.781) | 0.813 (0.809-0.818) | **0.820 (0.816-0.825)** |
| AUC at day 1 (95% CI) | 0.827 (0.820-0.834) | 0.795 (0.787-0.802) | 0.797 (0.789-0.805) | 0.828 (0.82-0.835) | **0.830 (0.822-0.837)** |
| AUC at day 2 (95% CI) | **0.807 (0.798-0.816)** | 0.752 (0.74-0.762) | 0.752 (0.742-0.762) | 0.800 (0.789-0.809) | 0.804 (0.795-0.814) |
| **Classification: overall length of stay < 14 days** |  |  |  |  |  |
| AUC (95% CI) | **0.826 (0.820-0.833)** | 0.777 (0.770-0.785) | 0.777 (0.770-0.784) | 0.820 (0.813-0.827) | 0.794 (0.797-0.802) |
| AUC at day 1 (95% CI) | **0.839 (0.828-0.85)** | 0.794 (0.782-0.808) | 0.794 (0.781-0.806) | 0.831 (0.818-0.843) | 0.809 (0.796-0.821) |
| AUC at day 2 (95% CI) | **0.826 (0.812-0.839)** | 0.782 (0.766-0.797) | 0.779 (0.764-0.794) | 0.815 (0.802-0.829) | 0.796 (0.782-0.809) |

**Table 13:** Summary of the results on predicting discharge destination for logistic regression (LR), CART decision trees (CART),optimal trees with parallel splits (OT), random forest (RF) and gradient boosted trees (GBT).

| Discharge Destination |  | LR | CART | OT | RF | GBT |
| --- | --- | --- | --- | --- | --- | --- |
| Weighted AUC (95% CI) |  | 0.561 (0.558-0.565) | 0.781 (0.779-0.784) | 0.769 (0.767-0.772) | **0.837 (0.835-0.839)** | **0.837 (0.835-0.840)** |
| Weighted AUC at day 1 (95% CI) |  | 0.583 (0.576-0.589) | 0.792 (0.785-0.798) | 0.764 (0.757-0.771) | **0.843 (0.838-0.849)** | 0.842 (0.835-0.847) |
| Weighted AUC at day 2 (95% CI) |  | 0.597 (0.589-0.604) | 0.781 (0.774-0.79) | 0.764 (0.757-0.772) | 0.838 (0.832-0.845) | **0.839 (0.832-0.845)** |
| Mortality (with DNR indicator) |  |  |  |  |  |  |
| AUC (95% CI) |  | 0.934 (0.929-0.938) | 0.946 (0.941-0.951) | 0.940 (0.935-0.945) | **0.962 (0.959-0.964)** | 0.951 (0.947-0.955) |
| AUC at day 1 (95% CI) |  | 0.952 (0.936-0.964) | 0.944 (0.927-0.96) | 0.949 (0.934-0.962) | **0.966 (0.957-0.974)** | 0.950 (0.934-0.966) |
| AUC at day 2 (95% CI) |  | 0.952 (0.934-0.965) | 0.947 (0.928-0.963) | 0.947 (0.93-0.963) | **0.965 (0.954-0.974)** | 0.949 (0.93-0.964) |
| Mortality (without DNR indicator) |  |  |  |  |  |  |
| AUROC (95% CI) |  | 0.855 (0.848-0.862) | 0.836 (0.829-0.843) | 0.838 (0.83-0.846) | **0.905 (0.900-0.909)** | 0.848 (0.84-0.856) |
| AUROC at day 1 (95% CI) |  | 0.904 (0.887-0.918) | 0.862 (0.84-0.882) | 0.862 (0.839-0.883) | **0.917 (0.901-0.93)** | 0.878 (0.855-0.898) |
| AUROC at day 2 (95% CI) |  | 0.900 (0.878-0.917) | 0.874 (0.851-0.895) | 0.858 (0.832-0.883) | **0.916 (0.898-0.931)** | 0.881 (0.857-0.903) |
| Extended Care Facility |  |  |  |  |  |  |
| AUC (95% CI) |  | 0.832 (0.829-0.835) | 0.804 (0.801-0.807) | 0.800 (0.797-0.803) | 0.850 (0.847-0.853) | **0.855 (0.852-0.858)** |
| AUC at day 1 (95% CI) |  | 0.858 (0.85-0.866) | 0.81 (0.801-0.819) | 0.805 (0.795-0.815) | 0.869 (0.862-0.876) | **0.873 (0.866-0.88)** |
| AUC at day 2 (95% CI) |  | 0.846 (0.837-0.854) | 0.803 (0.793-0.812) | 0.806 (0.797-0.816) | 0.861 (0.852-0.869) | **0.868 (0.86-0.876)** |

## D From individual risk scores to hospital-level estimates

In this supplementary section, we detail the two-step procedure we apply to convert individual probabilities for each patient to be discharged into estimates of discharge volume at a hospital level.

For each patient *i*, a machine learning model provides a score (between 0 and 1). Intuitively, this score constitutes an estimate of the probability for patient *i* to be discharged today, denoted 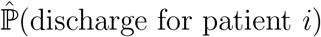. Consequently, by summing over all current inpatients, 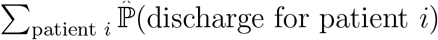 should be an unbiased estimator of the total expected number of discharges at the hospital. Unfortunately, depending on the machine learning method used and due to imbalance in the classes, the returned score might not be an unbiased estimate of the probability of discharge. Consequently, we use

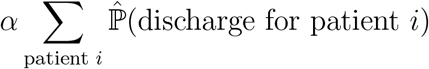

as an estimate of the number of discharges, where α is a correction factor, calibrated on the training set as the relative difference between the observed number of discharges and 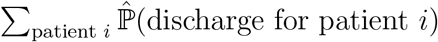 as in Van Walraven and Forster (2017).

Competitively, one can apply a “threshold-then-sum” approach where each patient is predicted as a discharge if her score exceeds some threshold *t* and then compute the expected number of discharges as the sum of predicted discharged patients, i.e.,

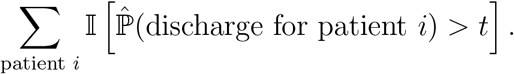

To obtain an unbiased estimate, *t* is chosen so that the number of false positives and false negatives on the training set are equal^3^.

We compare empirically these different aggregation strategies and report median absolute error in the daily number of discharges in Table 14. We make three observations out of these results: First, summing the raw scores returned by the machine learning models lead to useless estimates on the total number of discharges. Indeed, to handle class imbalance, observations are weighted according to their class prevalence. Weighting creates bias in the risk probabilities, bias which affects the training loss so as to be less sensitive to the imbalance in labels. However, once the model is trained, output scores conserve this bias and should be thus used carefully. A correcting factor or a “threshold-then-sum” approach is needed. Secondly, the correcting factor approach seems globally more accurate than “threshold-then-sum”. Note that this observation depends on the machine learning technique and is specific to the predictive task at hand. Finally, we also explore strategies where the correcting factor α or the threshold parameter *t* depends on the day of the week. Indeed, discharge volume is strongly impacted by day of the week-discharges are usually low on Sundays and high on Mondays -but this variable is not a first-order predictive feature in our individual risk models. Therefore, we account for this dependency in the aggregation strategy and obtained significant improvement, especially for the correction factor strategy. We use the latter in our implementation.

**Table 14:** Median Absolute Error in the daily number of discharges for logistic regression (LR), CART decision trees (CART), optimal trees with parallel splits (OT), random forest (RF) and gradient boosted trees (GBT), for various aggregation strategies.

| Aggregation method | LR | CART | OT | RF | GBT |
| --- | --- | --- | --- | --- | --- |
| $\sum_{\text{patient } i} \hat{\mathbb{P}}(\text{discharge for patient } i)$ | 99.9 | 109.7 | 10.2 | 20.5 | 7.8 |
| $\alpha \sum_{\text{patient } i} \hat{\mathbb{P}}(\text{discharge for patient } i)$ | 10.8 | 13.1 | 11.2 | 9.1 | 8.0 |
| $\sum_{\text{patient } i} \mathbb{I} \left[ \hat{\mathbb{P}}(\text{discharge for patient } i) > t \right]$ | 16.0 | 18.0 | 16.0 | 76.5 | 8.5 |
| $\alpha_{dow} \sum_{\text{patient } i} \hat{\mathbb{P}}(\text{discharge for patient } i)$ | 8.6 | 6.0 | 6.4 | 6.2 | 7.8 |
| $\sum_{\text{patient } i} \mathbb{I} \left[ \hat{\mathbb{P}}(\text{discharge for patient } i) > t_{dow} \right]$ | 14.5 | 17.0 | 15.0 | 76.5 | 10.5 |

## Footnotes

1 For ease of implementation and user-friendliness of the commercial package, we used OT trees over CART in the actual implementation.

2 In Table 7, we compare the output from the machine learning model with the middle of the interval provided by each nurse.

3 Note that this is not the convention used by the commercial implementation of Optimal Classification Trees (Interpretable AI 2020). As depicted in Figures 1-2, they choose t equal to empirical proportion of positive cases on the training population.

## Notes

### Competing Interest Statement

The authors have declared no competing interest.

### Funding Statement

No external funding was received to conduct the study

### Author Declarations

Institutional review board at BIDMC approved the study with waiver of informed consent.

### Summary of Updates

Author affiliations updated Empirical evaluation in Section 4.3 more robust and more clearly reported

